# Calibration-free regional RF shims for MR spectroscopy

**DOI:** 10.1101/2020.07.24.20161141

**Authors:** Adam Berrington, Michal Považan, Christopher Mirfin, Stephen Bawden, Young Woo Park, Daniel C. Marsh, Richard Bowtell, Penny Gowland

**Affiliations:** Sir Peter Mansfield Imaging Centre, School of Physics and Astronomy, University of Nottingham, Nottingham, UK; Russell H. Morgan Department of Radiology, Johns Hopkins University School of Medicine, Baltimore, MD, USA; F.M. Kirby Research Center for Functional Brain Imaging, Kennedy Krieger Institute, Baltimore, MD, USA; Center for Magnetic Resonance Research, Department of Radiology, University of Minnesota Medical School, Minneapolis, Minnesota, USA

**Keywords:** MR spectroscopy, parallel transmission, RF shimming, universal, 7 T, B_1_^+^

## Abstract

**Purpose:** Sufficient control of the RF transmit field (B_1_^+^) in small regions-of-interest (ROIs) is critical for single voxel MR spectroscopy at ultra-high field. Static RF shimming, using parallel transmit (pTx), can improve B_1_^+^, but must be calibrated for each participant and ROI, which limits its applicability. Additionally, specific-absorption-rate (SAR) becomes hard to predict. This work aimed to find RF shims, which can be applied to any participant, to produce the desired |B_1_^+^| within pre-defined target ROIs.

**Methods:** RF shims were found offline by joint-optimisation on a database, comprising B_1_^+^ maps from 11 subjects, considering ROIs in occipital cortex, hippocampus and posterior-cingulate, as well as the whole brain. The B_1_^+^ magnitude achieved using calibration-free RF shims was compared to a tailored shimming approach, and MR spectra were acquired using tailored and calibration-free RF shimming in 4 participants. Global and local 10g SAR deposition were modelled.

**Results:** Calibration-free RF shims resulted in similar |B_1_^+^| in small ROIs compared to tailored shimming, in addition to producing spectra of excellent quality and equivalent SNR. Only a small database size was required. SAR deposition was reduced compared to operating in quadrature mode for all ROIs.

**Conclusion:** This work demonstrates that static RF shims, optimised offline for small regions in single voxel MRS, avoid the need for lengthy B_1_^+^ mapping and pTx optimisation for each ROI and participant. Furthermore, power settings may be increased when using calibration-free shims to better take advantage of the flexibility provided by RF shimming for regional acquisition at ultra-high field.

## Introduction

MR acquisition at ultra-high field strengths (≥7T) is challenging, largely because of the inhomogeneity in the RF transmit field (B_1_^+^). This inhomogeneity arises from the interaction of the relatively short-wavelength RF with the sample, leading to spatial variation in the achieved B_1_^+^, which in turn causes spatial variations in image contrast^1,2^. The relative reduction of peripheral B_1_^+^ signal when using a volume RF coil leads to the commonly observed central brightening in the brain. For MR spectroscopy (MRS) using small regions-of-interest (ROIs), a lack of regional B_1_^+^ can necessitate higher transmit powers or longer duration pulses, thereby prolonging minimum TR or TE. Misadjusted B_1_^+^ leads to sub-optimal flip-angles, poor localisation and lower signal-to-noise ratio^3^. Thus, increasing the available B_1_^+^ in such regions is critical to obtaining the high spectral quality which is needed to take full advantage of the higher intrinsic signal-to-noise ratio and spectral resolution at ultra-high field^4,5^.

Parallel transmit (pTx) is now available on many MR systems and provides a means to control the B_1_^+^ profile across a sample^6–8^. B_1_^+^ can be statically ‘shimmed’ by tailored weighting of RF phase and amplitude on individual transmit channels. Static RF shimming can be performed using different strategies, for instance, by requiring that the mean transmit phases add constructively in the ROI^9,10^, or by maximising B_1_^+^ efficiency for the input power^11^, or by minimising variation in B_1_^+^ magnitude across the ROI using least squares optimisation^8,12^. Use of RF shimming in the brain has been demonstrated in multi-slice MRSI acquisition^13^ and in single voxel MRS^10^, for example. In the latter work, Emir *et al*. showed that RF shimming provides high spectral quality and improved quantification of metabolites from several ROIs. For instance, a 2.5-fold increase in the available B_1_^+^ magnitude was reported in the occipital region. In practice, RF shimming involves calibration and numerical optimisation for each ROI and each participant; mapping of the B_1_^+^ produced by each transmit channel is needed, as well as the manual definition of a target ROI. This process is time consuming (~10 minutes per ROI), which limits the use of RF shimming in practice, particularly for clinical studies and when multiple regions are scanned in a single session. Importantly, RF shimming also leads to significant changes in the electric field distribution and, hence, changes in SAR deposition.. Without an ability to monitor or predict SAR deposition, conservative limits are often placed on the allowable transmit power during RF shimming in order to avoid generation of high local SAR, and this further diminishes the benefits of RF shimming.

To avoid lengthy pTx calibrations, Gras et al. have proposed the use of ‘universal’ *k*_*T*_-points RF pulses for dynamic pTx^14^. These pulses were optimised offline over a database of acquired B_1_^+^ maps yet produced similar performance to pulses calculated on a participant-specific basis and required no B_1_^+^ mapping and numerical optimisation on the scanner. This approach relies on the similarity in B_1_^+^ profiles across participants and has led to development of calibration-free pTx refocussing pulses^15^ and selective local excitation pulses^16^. The calibration-free principle has also been extended using machine learning, where a set of RF pulses was calculated based on clusters of similar participants which better account for inter-subject variability^17^. To avoid extensive B_1_^+^ mapping, Ianni et al. developed a machine learning approach for whole-brain, slice-wise static shimming requiring only minimal B_1_^+^ information^18^.

RF shimming works best when applied to a small ROI^19^, thus it is expected that *static* shimming alone may be sufficient for regions examined with single-voxel MRS. Vendor-provided fixed amplitude and phase settings, calculated using electromagnetic simulations^22^, often cover large ROIs and may be sensitive to variations^20^ such as relative position or head size. Thus, in order to make the benefits of RF shimming more available for spectroscopic acquisition and given the universal pTx approach^14^, we hypothesised that jointly-optimising RF shim solutions over a database could generate shim weights that can achieve similar target B_1_^+^ magnitudes, compared to a tailored shimming approach, over a set of small pre-defined ROIs in any participant. Use of such an approach would account for natural variation across participants and would not require acquisition of B_1_^+^ maps for each participant.

In this work, therefore, calibration-free shim settings were calculated for a set of ROIs in commonly investigated brain regions (the occipital cortex, hippocampus and posterior cingulate), as well as the whole brain, using a database of B_1_^+^ maps from 11 subjects. ROIs are registered to the same location across participants using template definitions. The calibration-free RF shims were calculated offline, for a fixed total power transmitted through the coil elements. Results were compared *in vivo* to those produced by using tailored shimming in each case. MRS data were also acquired using the optimised shim settings on participants not included in the database. Finally, calibration-free RF shims were analysed in terms of their predicted SAR deposition compared to acquiring with the coil elements driven quadrature mode for a given ROI.

### Theory

#### RF shimming

The goal of this work was to generate static RF shim solutions, suitable for MRS, which reliably produce a desired magnitude of the RF transmit field (|B_1_^+^|) within an ROI without the need for B1 mapping and shim calculation on each individual participant. A magnitude-least-squares (MLS) optimisation was chosen as the RF shimming strategy. This minimises the mean squared difference between the shimmed |B_1_^+^| and a target value over a discrete set of spatial positions covering the ROI. This MLS strategy ensures that the mean |B_1_^+^| in the ROI approaches a target value while inhomogeneity is minimised.

RF shimming strategies typically maximise the |B_1_^+^| field in a region, within constraints on the maximum transmit power, and then renormalize the solution to obtain the desired flip-angle. In contrast, in this work RF shim solutions were constrained such that the sum of the power over all channel elements was limited *a priori* to ensure that the sequence will run within (vendor provided) safety limits on the maximum power per channel and for the required pulse shapes and sequence timings. This approach has the advantage that the same shim solution can be applied to every participant, using identical power settings. In addition, the |B_1_^+^| achieved using different shim solutions can be directly compared at the same fixed total power^8^. Obtaining more efficient shim solutions, which yield maximal |B_1_^+^| per unit power, requires flexible power constraints based on accurate SAR estimation and sequence-specific parameters.

In the following, |B_1_^+^| is expressed as a ratio of the value measured in the ROI (using B_1_^+^ mapping) to the prescribed value (i.e. the |B_1_^+^| required to achieve a particular flip-angle for a particular amplitude modulated pulse shape; specifically the nominal flip angle of the B_1_^+^ mapping sequence). Thus, |B_1_^+^| = 1 corresponds to achieving the desired B_1_^+^ in the ROI.

For each ROI, comprised of *N*_*ROI*_ discretised points (or image voxels), the RF shim phases and amplitudes applied to each transmit channel can be expressed as a complex *N*_*C*_ × 1 shim vector, *w*, where *N*_*C*_ is the number of transmit channels. The complex-valued B_1_^+^ in the ROI is expressed as an *N*_*ROI*_ × *N*_*C*_ transmit sensitivity matrix, *S*, such that the *i*^*th*^ row, *s*_*i*_ is the concatenation of B_1_^+^ values at voxel, *i* from all channels. Tailored shimming, performed on a participant-wise basis, involves minimising the MLS between the predicted and target |B_1_^+^| over the ROI, i.e. find

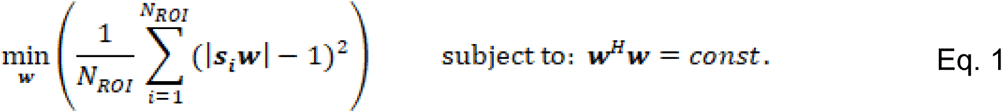

RF shim weights are calculated subject to a fixed total power, which is equal to that of quadrature mode where individual channel amplitudes are equal to unity (so ***w***^*H*^***w*** = 8, for an 8 channel system^8^).

#### Joint optimisation over the database

Similar to the formulation for universal *k*_*T*_-points pulses^14^, calibration-free shims were found for each ROI by calculating the shim vector, ***w***, which, when applied to each subject, *k*, in a database of K subjects, minimises the error for the worst-case subject in a minimax approach, i.e. find

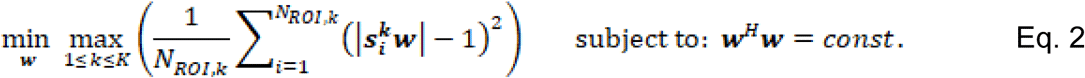

## Methods

### MR scanning

Data were acquired on a 7 T Philips Achieva MR system with a commercial pTx head coil (8Tx/32Rx, Nova Medical, Fig. 1A). A total of 15 healthy volunteers participated in this study with informed consent and with the approval of a local ethics committee. For all volunteers, the scanning protocol consisted of a T_1_-weighted MPRAGE acquisition (1 mm isotropic, TR/TE = 7.1/2.9 ms, TI = 1.1 s, TA = 4 m 8 s) and whole-brain B_1_^+^ mapping using a 3D DREAM^21^ sequence (3.5 mm isotropic, STE flip-angle (*α*) = 60°, excitation flip-angle (*β*) = 7°, TR/TE_STE_/TE_FID_ = 2.6/1.1/1.5 ms, TA = 9 s) acquired transmitting on all channels simultaneously. To obtain complex B_1_^+^ information for each transmit channel, low flip-angle 3D gradient echo images (3.5mm isotropic, TR/TE=8/6.7 ms, FA=1°, TA =3 m 51) were also acquired while transmitting on each channel separately. The |B_1_^+^| on each channel was calculated by normalising the GRE data on each channel to the complex sum over all channels and then scaling to the |B_1_^+^| map acquired with DREAM^22^. Transmit phase on each channel was obtained by comparing the phase of the GRE data from each channel to the first channel. When participants were scanned, no special attention was paid to participant head size or position in the coil. Anatomical and B_1_^+^ transmit data from the first 11 volunteers formed the training database for the calculation of calibration-free shims (D1-D11). The subsequent 4 datasets formed the test set (T1-T4) and were used to validate the calculated calibration-free RF shim settings. Demographics of the database and test sets were (28±5 yrs, 5F) and (47±12 yrs, 2F), respectively.

**Figure 1:**
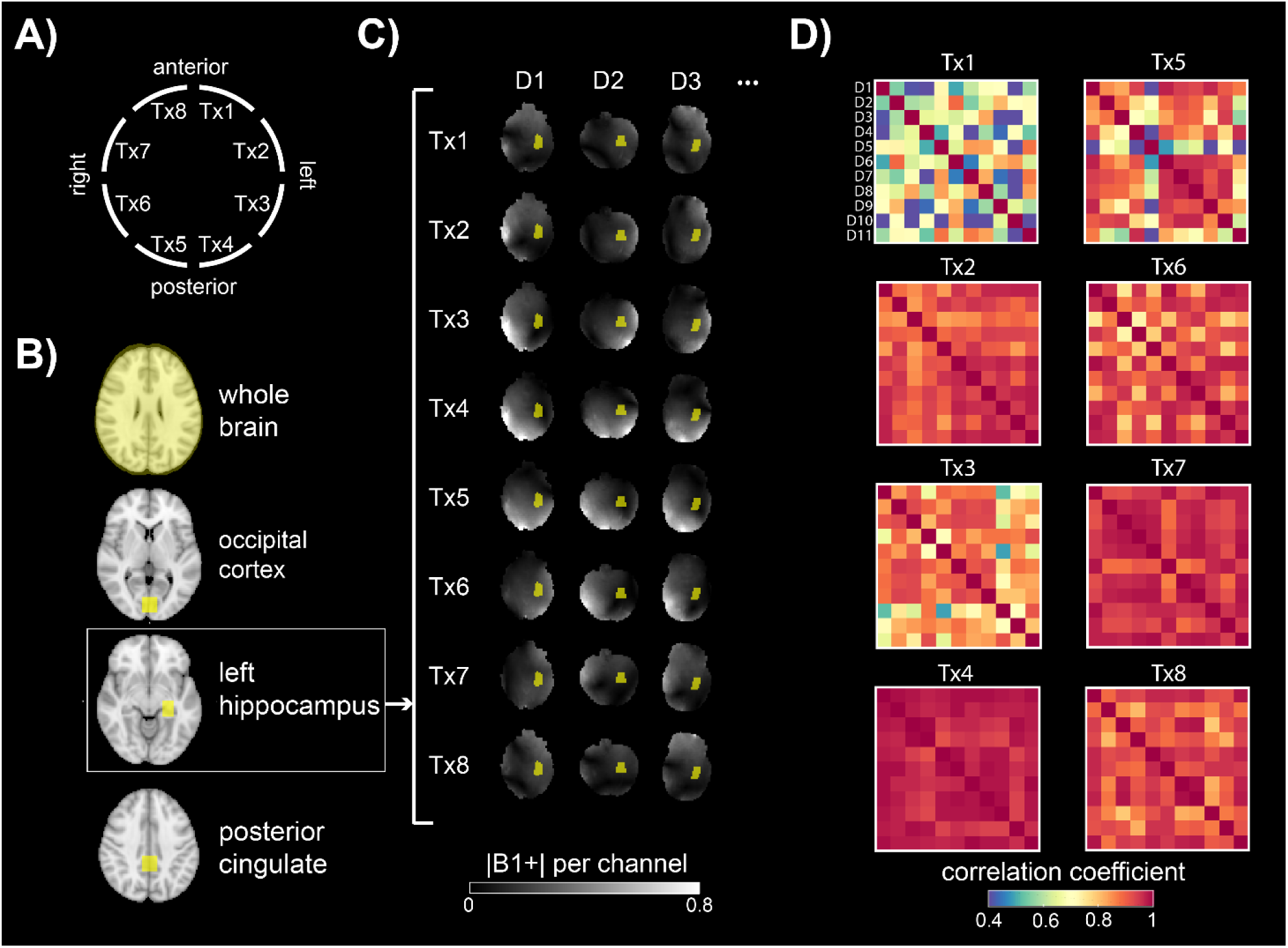
A) Schematic of the 8 transmit channels (Tx1-8) on the head coil. B) Pre-defined ROIs (in yellow overlay) defined in MNI space including 3 target ROIs for MRS (OCC, Hippo. and PCC). C) Registration of the hippocampal ROI from (B) into the participant’s |B1+| map for each transmit channel shown for three datasets from the database (D1-3). (D) B1+ correlation matrices for each transmit channel, showing the pairwise B1+ correlation between each subject in the database for the hippocampal ROI.

### ROI definition within the database

Target ROIs for 3 commonly measured regions in MR spectroscopy were defined on the MNI 1 mm brain atlas, namely, occipital cortex (OCC, 20×20×20 mm^3^), left hippocampus (Hippo, 30×15×12 mm^3^) and posterior cingulate cortex (PCC, 20×20×20 mm^3^), shown in Fig. 1B. In addition, a whole-brain ROI was defined. To obtain ROIs positioned in the same location in the B_1_^+^ maps of each participant, the T_1_-weighted images were registered to the lower resolution |B_1_^+^| maps acquired in quadrature with an affine transformation, and also to MNI template space using skull-stripping followed by a non-linear warp transformation (FSL^23^). ROIs were then transformed from MNI space, via the T_1_-weighted anatomical images, to the lower resolution set of 8 individual channel B_1_^+^ maps for each dataset (Fig. 1C) by applying the inverse of the successive transformations.

### Shim calculation

Tailored RF shimming at the console and offline joint-optimisation using the minimax approach for calibration-free shimming (Eq. 2) were performed using custom written code in MATLAB (The MathWorks, Natick, MA). Constrained optimisation was performed using the interior-point algorithm in *fmincon*. For tailored shimming on the scanner, the ROI for RF shimming extended beyond the MRS voxel by around 2 mm in each direction. Vendor-provided constraints on peak and average RF power were 2 kW and 1 W per channel, respectively. Average power was monitored on the scanner using vendor-provided power-monitoring software.

Shim weights from tailored and calibration-free shimming (Eq. 1 & 2) were scaled to that of the quadrature mode excitation. In quadrature mode the B_1_^+^ amplitude on each channel is the same (defined as unity, here) and the phase of each channel is incremented by 45° moving around the 8 elements on the cylindrical coil surface.

### B_1_^+^ correlation

To assess the degree of similarity in B_1_^+^ between each participant in the database, a correlational analysis was performed. B_1_^+^ maps for each channel were registered to MNI space and then masked by each ROI. For each transmit channel (Tx1-8), the complex B_1_^+^ distribution within the ROI was correlated between participants in a pixel-wise manner and the Pearson correlation calculated (Fig. 1D shows an example for the hippocampal ROI). As a summary statistic, the mean and standard deviations of these pairwise correlation coefficients averaged over all participants 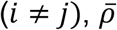, were calculated using Fisher transformation.

### MRS

For the test participants (T1-T4), MR spectroscopy data was additionally acquired in the three target ROIs using quadrature, tailored RF shimming and calibration-free shim weights. In addition, |B_1_^+^| maps were acquired using the RF shim settings. MRS was acquired using STEAM localisation and identical parameters for each target region (TE = 14 ms, TR = 4-5 s, number of transients (NT) = 48-64, pulse bandwidth: 5.7 kHz, pulse duration: 7.3 ms, peak B1: 15 μT, VAPOR water suppression^24^). Spectra acquired from all receive-channels were coil-combined, and then B0 and phase corrected before averaging. SNR was measured in the frequency domain as the height of the NAA peak divided by the standard deviation of the root-mean-square noise in the 10-12 ppm range.

### SAR modelling

SAR calculation for different shim modes was carried out using finite-difference time-domain electromagnetic simulations provided by Nova Medical, Inc. (computed with XFdtd, Remcom Inc., State College, PA). 3-dimensional vector magnetic (B1) and electric (E) field values were available for *Duke* and *Ella* dielectric head models computed on a 28×28×28 cm^3^ grid of resolution 2×2×2 mm^3^. Data were scaled using available scattering parameters, such that the forward power of each channel was 1 W in quadrature mode, which corresponded to an average |B_1_^+^| over the central transverse slice of 1 µT for both *Duke* and *Ella* models.

To calculate SAR for different RF shim settings, the Q-matrix approach was used^25,26^. Q-matrices were calculated at each position, *r*, using E-field, mass density and electrical conductivities^27^. Local SAR can then readily be computed by multiplication with the desired shim vector, *SAR*(*r*) = ***w***^*H*^***Q***(*r*)***w***. By averaging over all Q-matrices, a global Q-matrix, and hence mean head SAR, can be obtained for the whole model. Local 10g SAR was calculated at each position in the model. 10g cuboidal regions were found using an iterative approach^28^ and regions with a large proportion (>20%) of zero-mass voxels were rejected. Local SAR was then calculated using a weighted sum of the Q-matrices within each 10g region. SAR computations were carried out on a high-performance computing cluster (2×20 core processors and 192 GB RAM). The RF duty cycle was assumed to be 100%. SAR in (W/kg) was computed relative to the B_1_^+^ field measured in quadrature for both models.

### Cross validation

To estimate the effect of database size on the performance of the calibration-free RF shims, a cross-validation simulation was performed. The size of the database was incremented from n = 1 to 11. At each increment, calibration-free shims for every possible combination of n training data were calculated, and the resulting B_1_^+^ maps for 3 test datasets were evaluated. The |B_1_^+^| and RMSE were calculated and averaged over all combinations for each increment of database size.

## Results

### B_1_^+^ correlations across database participants

For each ROI, the pairwise correlation of B_1_^+^ across the ROIs between all (n=11) datasets in the database was measured for each transmit channel (Fig 1D). The mean values of these correlation coefficients, averaged across the 11 subjects in the database are shown for each ROI in Fig. 2. The B_1_^+^ over the whole brain ROI was well correlated between subjects for all transmit channels (min. 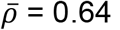 for Tx channel 4, max. 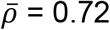 for Tx channel 7). When considering the three target ROIs, the mean B_1_^+^ correlation across the database participants was generally weakest within the occipital region (OCC) (Tx channel 2 gave minimum 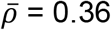; Tx channel 8 had highest 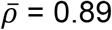). B_1_^+^ correlation within left hippocampus and posterior cingulate ROIs were higher than OCC and followed similar trends across channels; Hippo. (min. 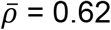on Tx1; max. 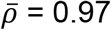 on Tx4) and PCC (min. 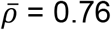on Tx7; max. 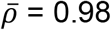on Tx6).

**Figure 2:**
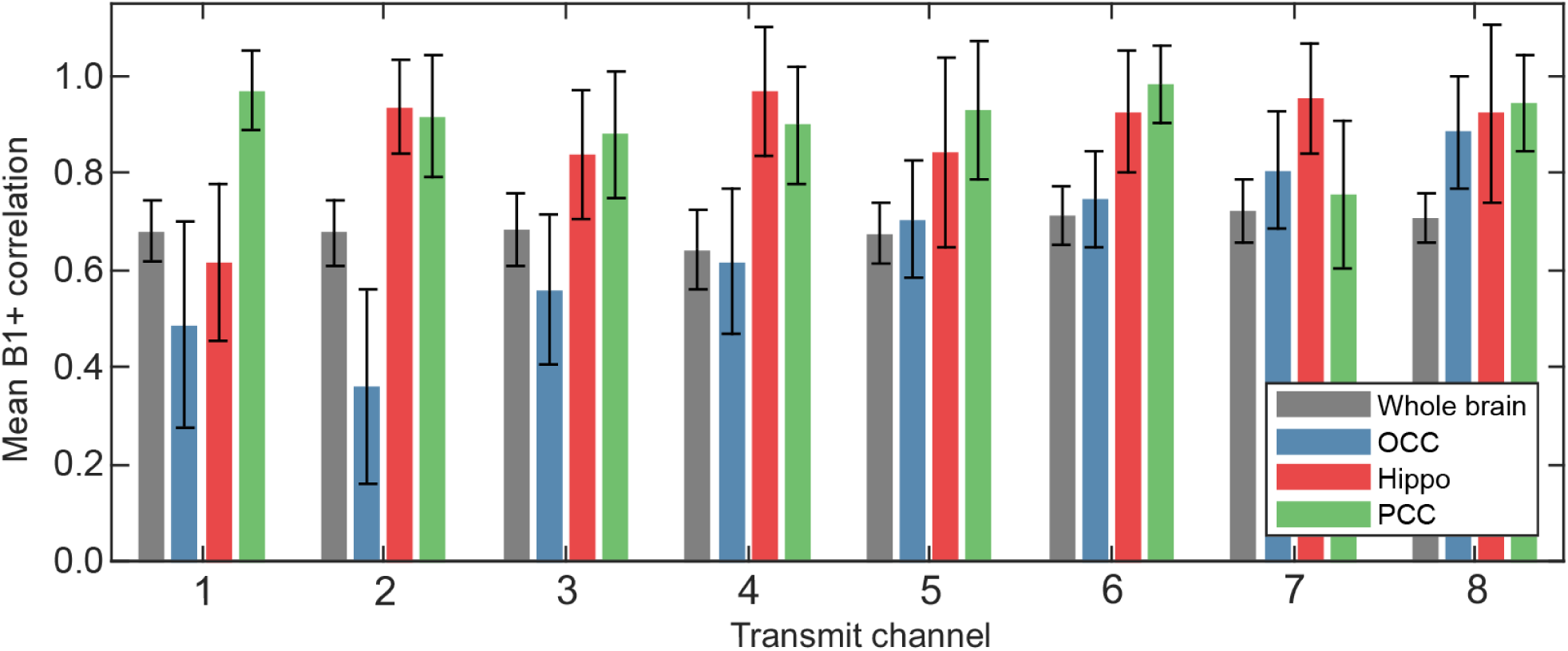
Mean regional correlation coefficients of B1+ on each channel across the 11 participants in the database (D1-11). Correlations determined for the whole brain and three regions (occipital, hippocampus and posterior cingulate).

### RF static shim solutions

The static RF shim weights arising from tailored shimming (Eq. 1) for the database participants (D1-D11) are shown on a polar plot in Fig. 3A for all 3 target ROIs. RF shim amplitudes are scaled relative to quadrature mode. Corresponding solutions for whole brain RF shimming are provided in Supporting Information A. For tailored shimming, OCC shims showed the greatest variation between database participants in channel phase (standard deviation: 95°, Tx8) and amplitude (standard deviation: 0.26, Tx1) of any target ROI.

**Figure 3:**
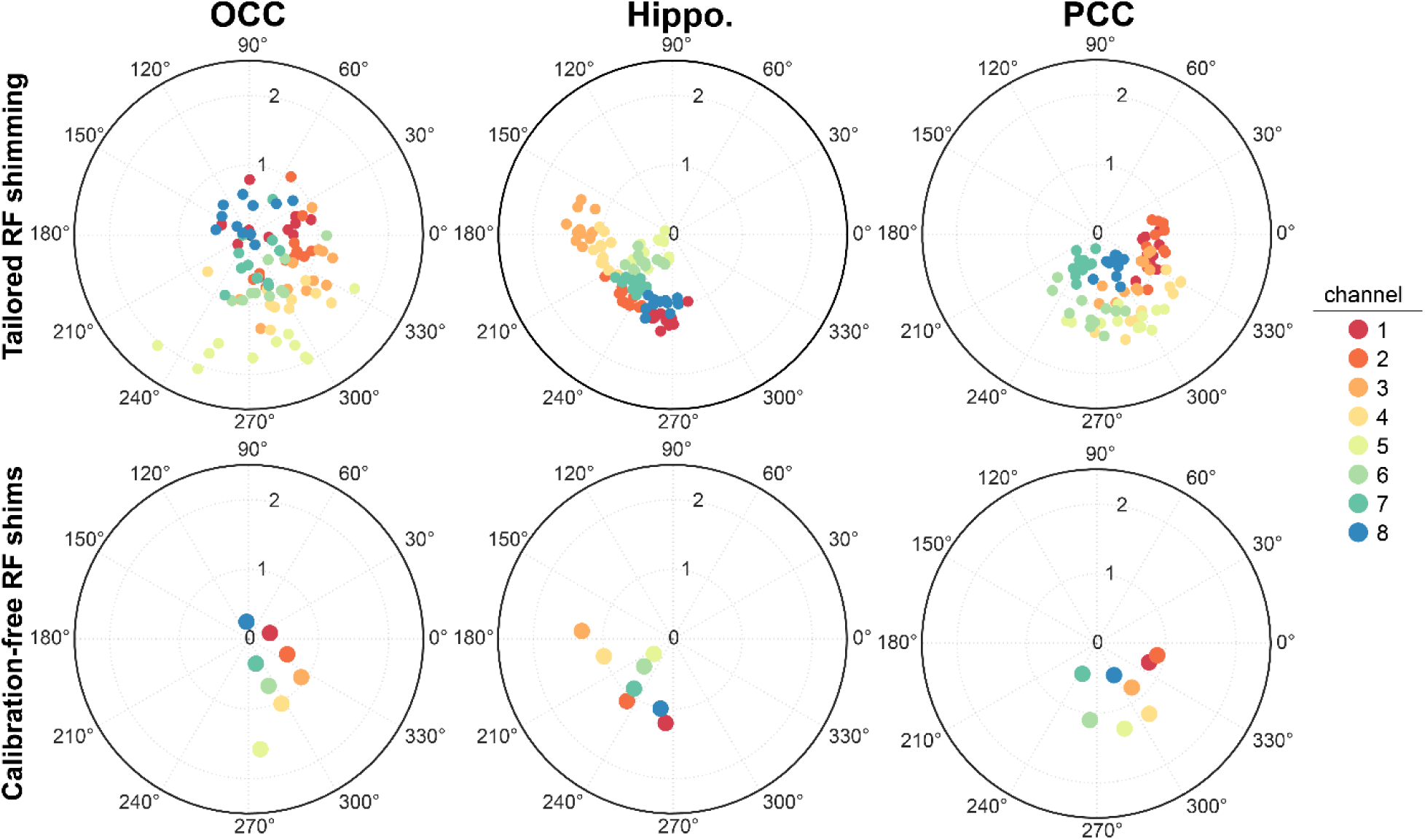
RF shims (phase and amplitude) on each transmit channel calculated for the three different ROIs when applying tailored shimming across B1+ maps in 11 different subjects (top) and for calibration-free shims produced by optimising over the whole database (bottom). RF shim amplitudes are scaled such that in quadrature mode the amplitude on each channel is 1.

The calibration-free static RF shims (Eq. 2), calculated over the database participants are shown in Fig. 3B for all 3 target ROIs. The distribution of channel phases for each ROI roughly matched that of tailored shimming (Fig 3A). OCC calibration-free shim weights had the largest single-channel amplitude (max |*w*_*i*_| = 1.64, Tx channel 5) and the mean relative phase between neighbouring transmit elements was 81°, 93° and 32° for OCC, Hippo and PCC, respectively.

### Predicted |B_1_^+^|

The predicted |B_1_^+^| was calculated in each ROI using the tailored and calibration-free shims and compared to the quadrature mode values (Fig. 4) for the database subjects (D1-D11). Calibration-free RF shims led to statistically similar |B_1_^+^| levels in all 3 target ROIs compared to tailored RF shimming. When comparing tailored to calibration-free shimming for the database subjects, there was no significant difference between mean |B_1_^+^| in the ROIs in OCC (0.99 vs. 0.96, p = 0.15), left hippocampus (0.97 vs. 0.96, p = 0.5) or PCC (0.93 vs. 0.93, p = 0.98). However, there was a difference in |B_1_^+^| in the whole-brain ROI for tailored compared to the calibration-free approach (0.73 vs. 0.66, p<1E-4).

**Figure 4:**
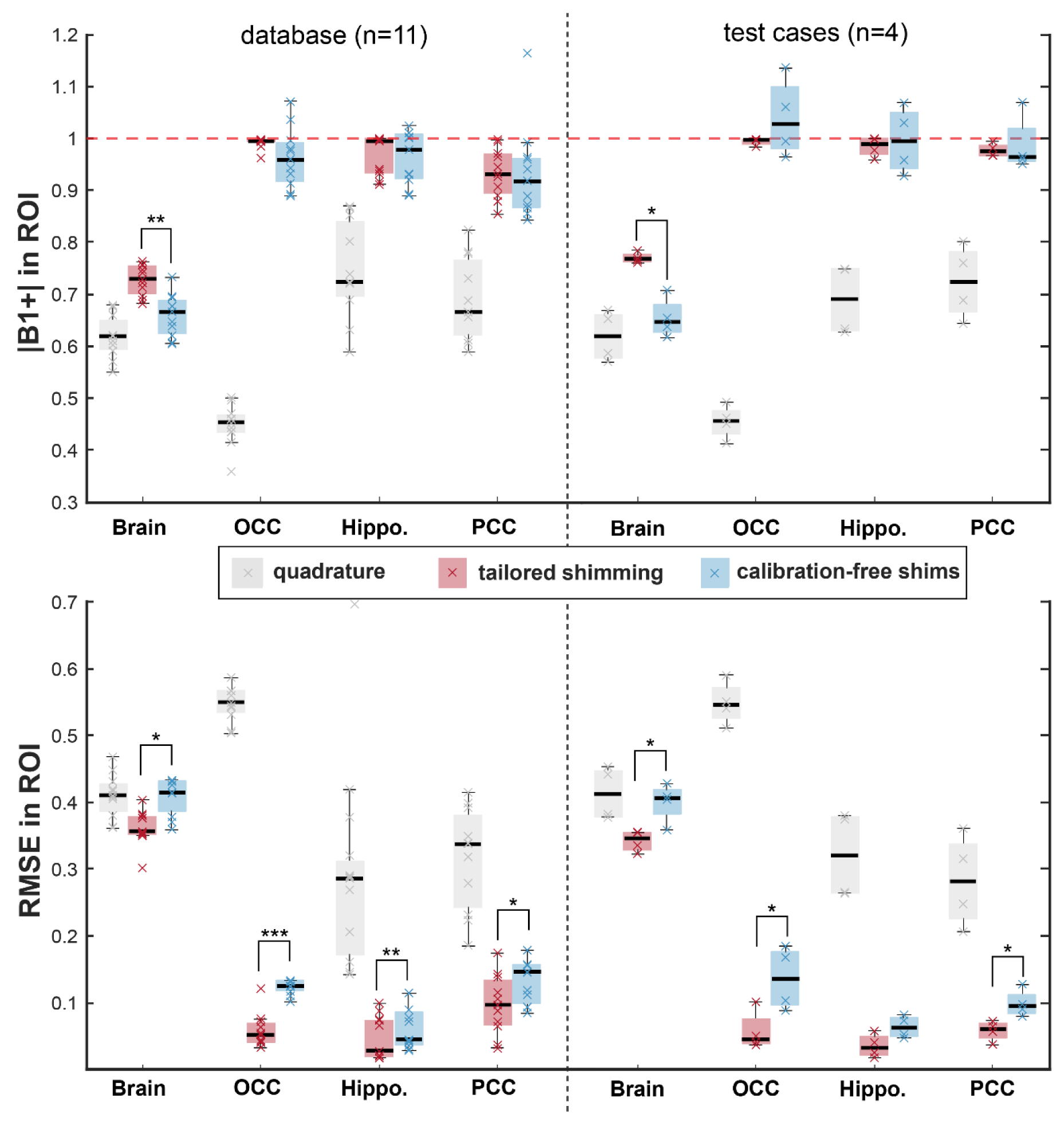
Predicted mean |B1+| and RMSE in all ROIs (Whole Brain, OCC, Hippo and PCC), calculated after relevant RF shim was applied (tailored or calibration-free). Analysis was performed on the database (left) and the test datasets (right). Red horizontal dotted line indicates the target value of |B1+|=1. RMSE and |B1+| were significantly different for all ROIs with RF shimming compared to quadrature mode (statistical comparison shown between tailored and calibration-free shims) (* p <0.05; ** p <1E-4, *** p < 1E-5). Lines through each bar indicate median value and shaded region represents 25^th^ - 75^th^ percentiles.

The RMSE of predicted |B_1_^+^| from the target over the ROIs are also plotted. There were small, but significant differences in RMSE in OCC (0.06 vs. 0.12, p < 1E-5), Hippo. (0.05 vs. 0.06, p < 1E-4) and PCC (0.10 vs. 0.13, p = 0.02) ROIs as well as whole brain (0.36 vs. 0.41, p <5E-4) when using tailored vs. calibration free shimming, suggesting an increased variability in |B_1_^+^| across the ROI for the calibration free case. As expected, RF shimming led to significantly reduced RMSEs and higher |B_1_^+^| in all three target ROIs compared to quadrature mode operation. For the whole-brain ROI, calibration-free shims did not lead to an improved RMSE compared to quadrature mode (p=0.94).

The same comparison was performed on the B_1_^+^ maps acquired in the test dataset (T1-T4), i.e. those participants who were not included in the database optimisation. After applying calibration-free shims, the mean |B_1_^+^| in the test dataset after calibration-free shimming was statistically similar to that obtained in the database subjects for all ROIs (OCC: 1.0, p =0.06; Hippo: 1.0, p=0.3; PCC: 0.99, p = 0.29; whole brain: 0.65, p=0.8). In addition, the RMSE was statistically similar to that obtained in the database subjects for all regions: RMSE: OCC (0.14, p=0.4), Hippo: (0.06, p=0.95), PCC: (0.10, p=0.09), whole brain (0.4, p=0.6).

Figure 5 shows |B_1_^+^| maps acquired in vivo in subjects T1-T4 using calibration-free and tailored shims for the PCC ROI, and also using quadrature mode operation.

**Figure 5:**
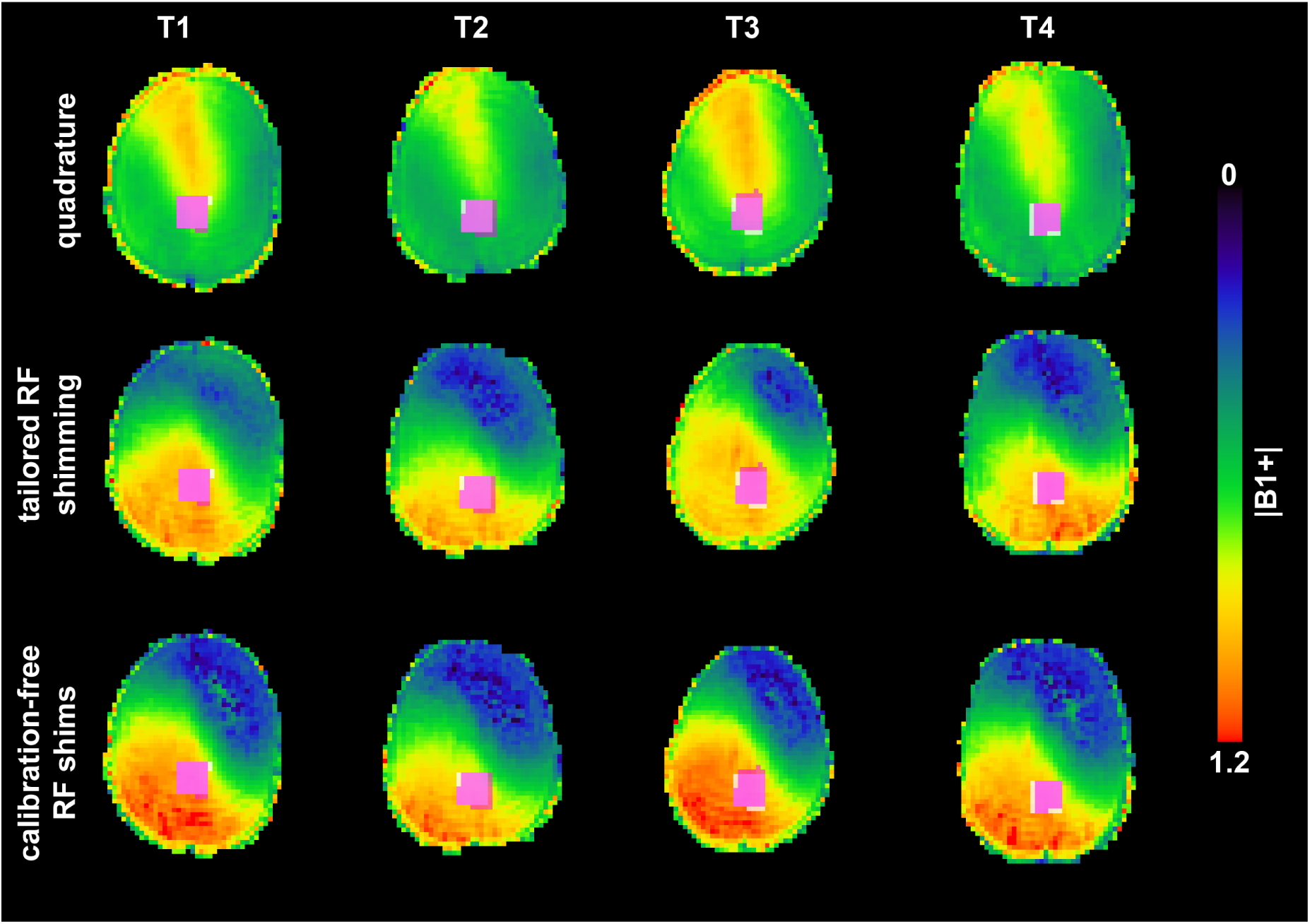
|B1+| maps (shown for a transverse slice) acquired from four test datasets (T1-4) outside of the database. Data was acquired with three different RF channel weightings for PCC acquisition. The final row is acquired with the calibration-free shims. Pink box is the calibration-free target ROI in B1+ space of each participant overlaid onto the MRS voxel position (white box). Images were skull-stripped.

Calibration-free shims generated a pattern of |B_1_^+^| inside the target ROI, and across the whole brain, that was reproducible across subjects (T1-T4), for all target ROIs.

### Spectral profile and SNR

Spectra from a representative test subject (T3) are shown in Figure 6 and spectra from other subjects are provided in Supporting Information B. Spectra acquired in all ROIs using calibration-free RF shimming were of excellent quality and all showed a small residual water peak. Spectral profiles for tailored and calibration-free shimming were remarkably similar even for the challenging hippocampal region. Compared to operating in quadrature mode, RF shimming led to cleaner spectral profiles with less lipid and outer voxel contamination. Using the power settings applied across shim modes in this work, it was not possible to obtain metabolite spectra from the OCC region in all test subjects when operating in quadrature mode (mean |B_1_^+^| = 0.45±0.04) due to a lack of signal for projection-based B0 shimming. The mean spectral SNR obtained with tailored RF shimming was not significantly different from that obtained with calibration-free shims; 119±6 vs. 122±15 in OCC (p=0.54), 50±3 vs. 49±3 in Hippo. (p=0.27) and 108±30 vs. 110±34 in PCC (p=0.86). Overall, there was no detectable difference in spectral quality between tailored and calibration-free shimming.

**Figure 6:**
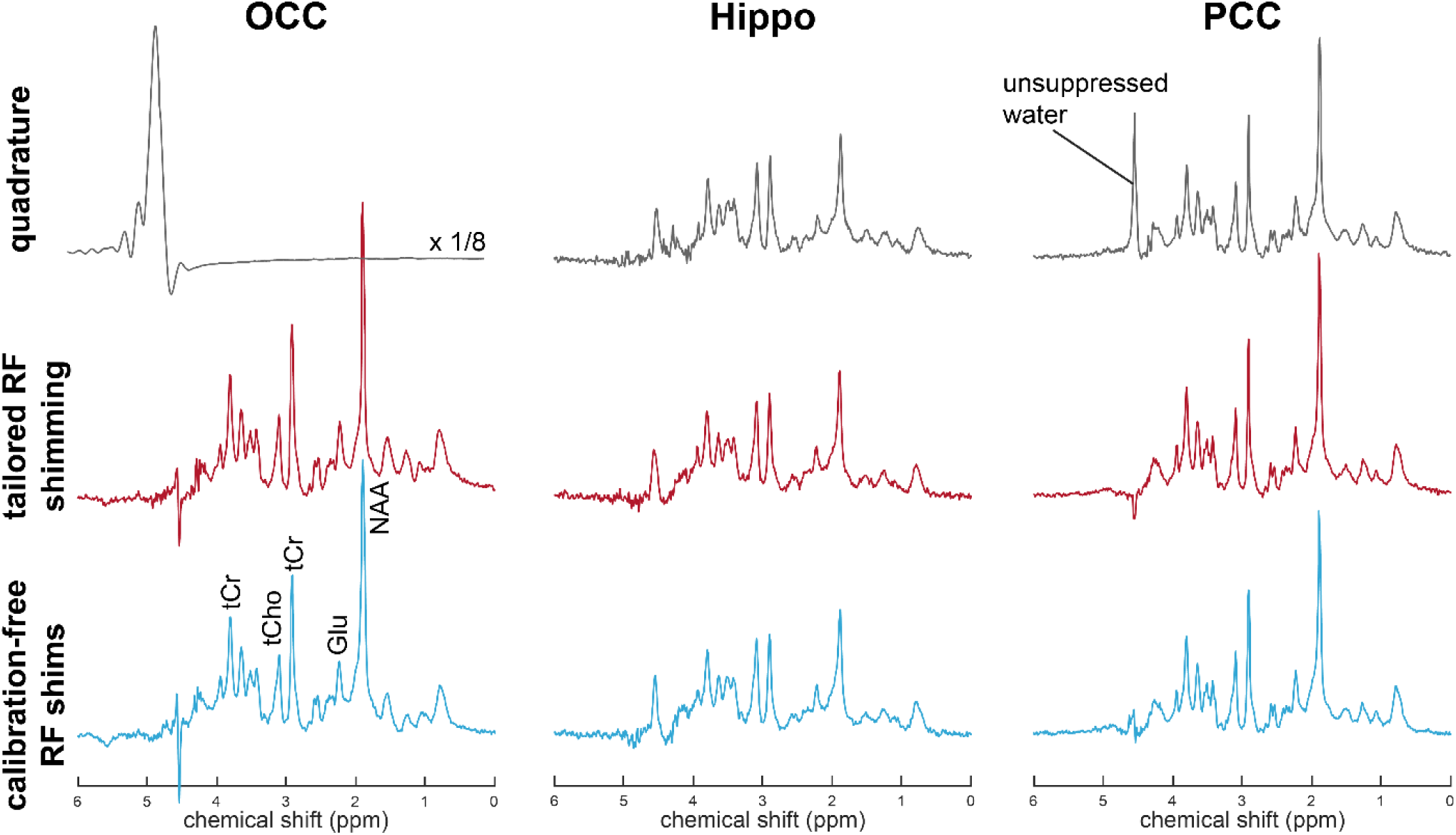
Spectra acquired in one subject (T3) from the three ROIs using quadrature mode operation and tailored and calibration-free RF shim settings. Spectra were Gaussian filtered (σ^2^=0.2) for display only.

### Effect of size of database

The influence of the size of the database used in calculating the calibration-free shims was estimated using a cross-validation approach and the results are shown in Figure 7 for the three target ROIs. As the number of datasets for calibration-free shim calculation increased from 1 to 11, the mean |B_1_^+^| remained close to the target value even with few available datasets. The mean RMSE in all target ROIs appeared to plateau. There was a consistent reduction in RMSE for the OCC target ROI with inclusion of up to 6 datasets, beyond which the gains in RMSE diminished. The PCC and hippocampal ROIs, however, did not show a decrease in RMSE with increasing number of datasets. In fact, the inclusion of only a single dataset was sufficient to reach a stable RMSE value in these ROIs.

**Figure 7:**
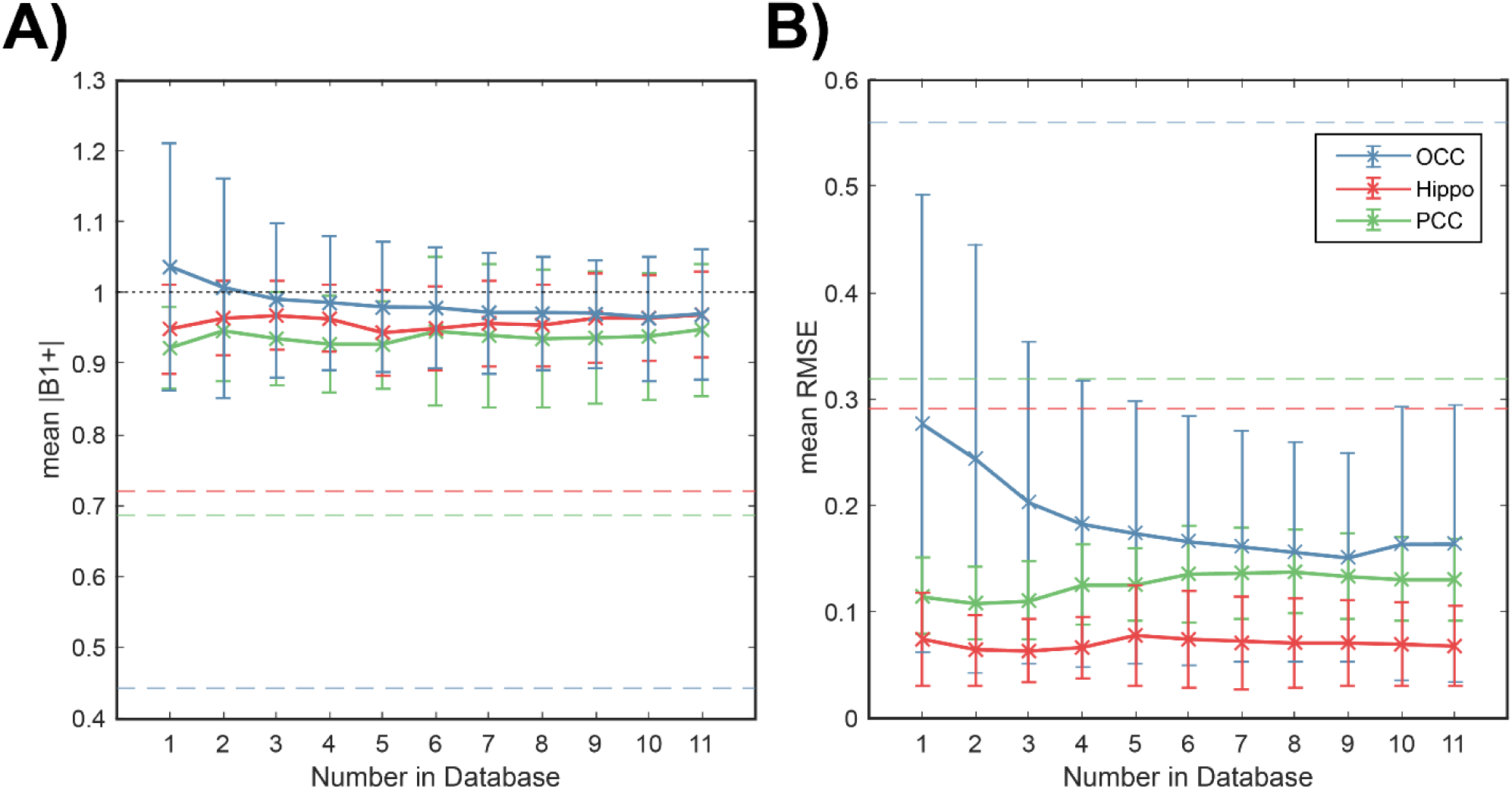
A) Mean |B1+| and B) mean RMSE after the calculation and application of calibration-free RF shims with increasing number of B1+ maps in the database. The simulation was carried out as a cross validation from a total of 15 available B1 maps, with each iteration using 3 test datasets. Dashed line indicates average RMSE in region when run in quadrature mode across the simulated datasets. Black dotted line is target |B1+|=1.

### SAR

Simulations of SAR deposition using the calibration-free shim settings were carried out on standard dielectric head models, Duke and Ella. Fig. 8 shows a map of the average local 10g SAR for both models after the hippocampal ROI calibration-free RF shims were applied with results scaled to the same total transmitted power as in quadrature mode. The pattern and location of hotspots was similar for both dielectric models. Maximum local 10g SAR using these shim settings was closely matched between models: 3.79 and 3.74 W/kg for Duke and Ella, respectively. This was lower than respective values obtained in quadrature mode by 84% (4.51 W/kg) and 75% (4.99 W/kg). SAR maps for the Hippo. and PCC ROIs are provided in Supporting Information C.

**Figure 8:**
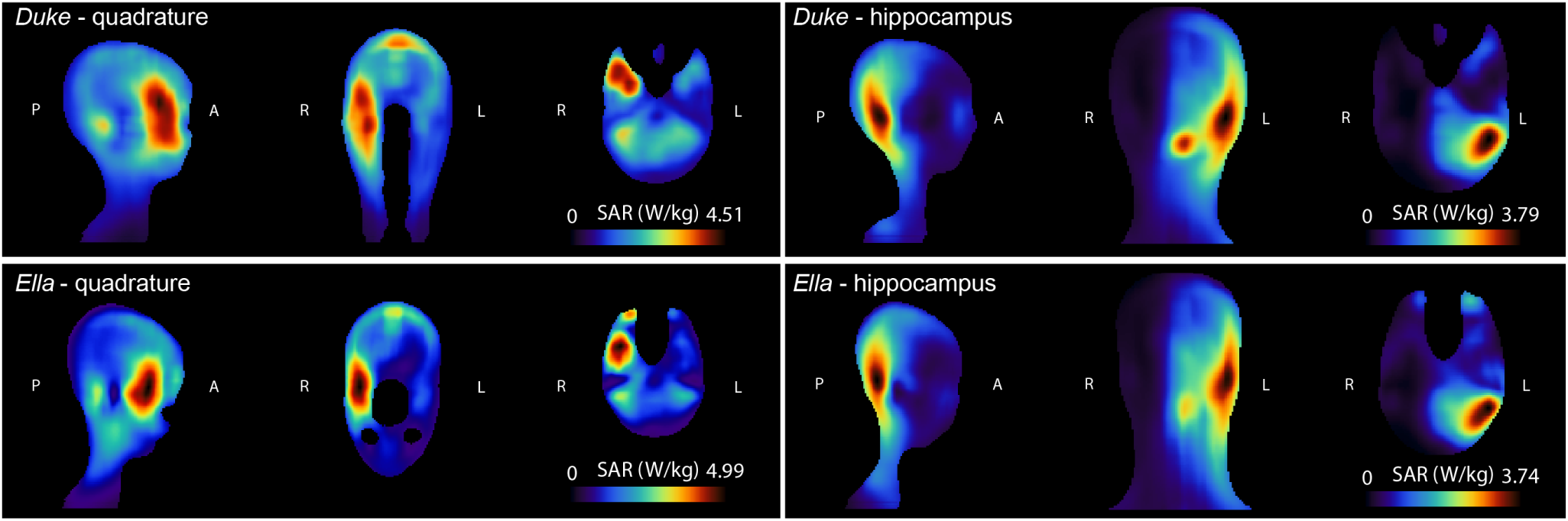
Average local 10g SAR (W/kg) simulated for Duke and Ella dielectric head models after the calibration-free RF shim for the hippocampal ROI was applied. RF shim weights for SAR estimation are normalised to generate a |B_1_^+^| of 1uT in quadrature mode. The sagittal, coronal and axial slices shown intersect at the maximum local 10g SAR (Duke: 3.79 W/kg and Ella: 3.74 W/kg).

A summary of the SAR results is given in Table 1. Average head SAR deposition using calibration-free shims was less than in quadrature mode for both models with a maximum average head SAR of 0.93 W/kg for OCC in Ella (65% of quadrature mode). The lowest total head SAR was observed in the Duke model using the Hippo. calibration-free shims (0.66 W/kg; 52% of quadrature). Across all ROIs, calibration-free shims produced lower maximum local 10g SAR deposition than quadrature mode. The PCC shims for Duke gave the highest 10g SAR (4.32 W/kg, 96% of quadrature) whilst the OCC shims for Ella gave the lowest (3.59 W/kg, 72% of quadrature).

**Table 1:** Summary of SAR simulations for four different RF settings (quadrature, OCC, Hippo., PCC) for Duke and Ella models. Head SAR was averaged over the whole model. Location is of maximum local 10g SAR and values are given relative to those obtained using quadrature mode excitation (% of Quad).

| Shim | Model | Head SAR (W/kg) | % of Quad | Max 10g SAR (W/kg) | % of Quad | Location (x,y,z) (image pixels) |
| --- | --- | --- | --- | --- | --- | --- |
| Quad | Duke | 1.27 | - | 4.51 | - | (90,85,54) |
|  | Ella | 1.44 | - | 4.99 | - | (92,79,66) |
| OCC | Duke | 0.82 | 65% | 3.55 | 79% | (72,59,88) |
|  | Ella | 0.93 | 65% | 3.59 | 72% | (65,32,63) |
| Hippo | Duke | 0.66 | 52% | 3.79 | 84% | (95,47,66) |
|  | Ella | 0.70 | 48% | 3.74 | 75% | (96,46,64) |
| PCC | Duke | 0.71 | 56% | 4.32 | 96% | (69,49,80) |
|  | Ella | 0.85 | 59% | 4.47 | 90% | (71,48,81) |

## Discussion

This work has demonstrated a method of acquiring B_1_^+^ calibration-free MRS data at 7 T with a commercial pTx coil by designing static RF shim solutions, for pre-defined regions of interest, generated from a database of B_1_^+^ maps and anatomical images. The |B_1_^+^| generated in small ROIs using calibration-free RF shims was not significantly different to that produced using subject-specific tailored RF shims, even in areas where large B_1_^+^ variation existed across database participants. The calibration-free shims produced consistent |B_1_^+^| for a given power setting so that renormalisation of the target flip-angle was not required. The approach requires neither B_1_^+^ mapping, nor prior definition of a B_1_^+^ shimming region, nor numerical optimisation of the shim settings at the scanner console, thereby alleviating the time-consuming steps of using pTx. Calibration-free RF shims led to spectra of excellent quality, with adequate water suppression and comparable SNR to that obtained using tailored RF shimming. By their nature, calibration-free RF shims allowed more predictable and easier offline SAR modelling than tailored shims and led to more consistent B_1_^+^ patterns across the brain. Reduced global and local 10g SAR was generated for the same total transmitted power when using the calibration-free shims compared to quadrature mode, suggesting that transmit power could be increased. It is anticipated that this method could be extended to any small ROI. Consequently, calibration-free static RF shims may increase the practical advantage of pTx for ultra-high field MRS, while cutting down scan time, to make such measurements more clinically viable.

Static RF shim solutions were sufficient to generate reproducible |B_1_^+^| profiles across participants in small target ROIs for single voxel MRS, yet mean |B_1_^+^| in the larger whole brain ROI was far below the desired value and was worse for calibration-free shims than tailored shimming. This is in line with the inferior performance of static RF shimming over whole brain ROIs in the work of Gras *et al*.^14^ and also with simulations performed by Mao *et al*. reporting that RF shimming is more effective for small ROIs^19^. Although mean pairwise B_1_^+^ correlation between database participants for the whole brain ROI in the current study was lower (0.64, worst case) than that of Gras *et al*. (0.82, worst case), calibration-free shim solutions could be readily found to produce comparable |B_1_^+^| availability in small ROIs to tailored RF shimming. The lower correlations in this study may be due to variation in head position relative to RF coil elements and head size of the participants, which was not controlled for as the database was formed. Considering B_1_^+^ correlations within the 3 target ROIs, the worst-case agreement was observed in the OCC region. The OCC also required more subjects in the database to reach a stable RMSE compared to the other two regions when calculating the optimal calibration-free shims (Fig. 7) and the OCC had a larger variation in RF shim phases and amplitudes when applying tailored shimming. As a result, OCC measurements showed the greatest benefit from B_1_^+^ shimming, similar to previous findings^10^. The OCC is the most inferior and peripheral of all the target ROIs studied, and showed the lowest |B_1_^+^| when running in quadrature mode due to the loss of peripheral B_1_^+^ from destructive interferences in head using volume transmit coils^2^. The OCC is closest to the coil elements and it is likely that participant head position in the coil varies in the coronal (L-R) and sagittal (A-P) planes, thus the relative position of the OCC with respect to the coil elements is presumably most variable.

Calibration-free shims were calculated with an *a priori* constraint on the total power summed across all transmit elements. The total power was fixed at a value, determined experimentally, such when applying RF shimming using the pulses and timings of the sequences used, the average power through any channel remained less than 1 W per channel (the vendor-determined average safety limit). By choosing the total power of the shim solution *a priori* based on safety limits for a given sequence, more efficient use of power is not possible and thus is a limitation of this work. A consequence of fixing the power for all acquisition modes to fall below vendor-provided safety limits for RF shimming, is that quadrature mode performance may have been improved by increasing the input power to achieve a desired |B_1_^+^|. Conversely, by relaxing the total power used in RF shimming, the target |B_1_^+^| in any ROI may have been reached using less power than presented here. An equal power setting across acquisition modes, however, allowed for accurate comparison of |B_1_^+^| and RMSE for the same input power. In general, the approach presented here is flexible and may be applied with different shimming strategies. In future, a softer constraint on total power during the shim optimisation may be more appropriate in addition to taking into account sequence-specific parameters to allow more efficient use of power^29^.

RF shimming strategies can produce lower global and local SAR than quadrature excitation as predicted from the targeted use of available B_1_^+^^27,30^. This finding was confirmed in the current work, where head SAR and local 10g SAR were considerably lower using calibration-free optimised RF shims compared to quadrature mode. The OCC calibration-free RF shims produced the highest head SAR (0.93 W/kg) out of all ROIs, yet lowest maximum local 10g SAR (3.55 W/kg). Interestingly, OCC shims had the largest single channel amplitude (Fig. 3). The smallest mean phase difference between neighbouring transmitter channels was for the PCC calibration-free shims, which also produced the highest local 10g SAR.

The spatial distribution of calculated head SAR using calibration-free shims was similar across models and local hotspots overlapped with the target ROI. This is important since SAR information is not readily available during a scan, and so conservative power settings are often employed when performing RF shimming^30^, as was the case in the current work. However, calibration-free shimming can be used in conjunction with offline SAR modelling to provide better estimates of local and global SAR allowing higher transmit power to be tolerated. Thus, the advantages of pTx may be more readily realised. Ideally, SAR variance would be calculated directly using models developed from database participants to determine power safety margins. In this work, however, that was not possible since precise coil models were not available.

The results for the whole brain ROI showed that as the size of the target ROIs increases, for example over a large MRSI grid, static RF shimming alone is not able to produce desirable solutions and the inclusion of RF and gradient waveform optimisation is required. However, the design of large flip-angle spokes pulses^31^, which have a broad frequency profile to reduce chemical shift displacement for MR spectroscopy is non-trivial^32^. Additional work is needed, therefore, to design broadband spokes pulses in order for the universal approach to be applied to larger ROIs in MR spectroscopy.

The use of a template definition for calibration-free regional shims requires that the measurement region overlaps with the template. In the current work, this was not an issue (Fig. 5), although it is anticipated that variability in voxel placement by scanner operators may be problematic for template-based ROIs. Automatic voxel placement has been presented as a solution to operator variability in single voxel MRS^33^ and would allow identical definitions of ROIs for RF shimming and placement, ensuring maximal overlap. Finally, it is expected that, as with universal *k*_*T*_-pulses, calibration-free static RF shim settings may be applicable across sites for the same coil and head position and the approach may be extended to different organs. Thus, a dictionary of shim settings for many commonly measured regions could be generated to simplify the improve repeatability in cross site and even cross vendor studies, using the same RF coil.

## Conclusion

Static RF shims calculated offline over a database of participants for pre-defined ROIs, produced high quality MR spectra at ultra-high field. Results were similar to those produced by performing tailored RF shimming on each participant and region separately, but the scanning time was reduced in the case of the calibration-free shims because B_1_^+^ mapping and on-line shim calculation was not required. The resulting SAR deposition from calibration-free shims was lower than in quadrature mode and, intrinsically more predictable than when tailoring the shimming for each participant, suggesting that such shims could be implemented using a higher power constraints than typically imposed on RF shimming in most pTx systems. As a result, the practical ease and utility of pTx for ultra-high field MRS studies is significantly improved using the calibration-free approach.

## Data Availability

Computational code relevant to the current manuscript will be made freely available and is available on request.

## Acknowledgements

The authors would like to acknowledge the support of the Precision Imaging Beacon and the Sir Peter Mansfield Imaging Centre, University of Nottingham. In addition, we particularly thank Andrew Peters for his management of the facilities as well as Peter van der Leuven from Philips Healthcare for discussion about RF safety limits, and NOVA Medical for providing FDTD simulations of the 8Tx32Rx coil. The SAR computations described in this paper were performed using the University of Nottingham’s Augusta HPC service, which provides a High Performance Computing service to the University’s research community.

